# Mental health status among family members of health care workers in Ningbo, China during the Coronavirus Disease 2019 (COVID-19) outbreak: a Cross-sectional Study

**DOI:** 10.1101/2020.03.13.20033290

**Authors:** Yuchen Ying, Fanqian Kong, Binbin Zhu, Yunxin Ji, Zhongze Lou, Liemin Ruan

**Affiliations:** Medical School of Ningbo University, Ningbo, Zhejiang 315010, P.R. China; Department of Psychosomatic Medicine, Ningbo First Hospital, Ningbo Hospital of Zhejiang University, Ningbo, Zhejiang 315211, P.R. China; Department of medical record and statistics, Ningbo Medical Center Lihuili Hospital, Ningbo,Zhejiang 315041, P.R.China; Department of anesthesiology, the Affiliated Hospital of Medical School of Ningbo University, Ningbo 315020, P.R. China

**Keywords:** Coronavirus Disease 2019 (COVID-19), mental health, family members of health care workers, generalized anxiety disorder, depressive symptoms

## Abstract

**Background:** So far, the psychological impact of COVID-19 epidemic among family members of Health care workers (HCWs) in China has been neglected. The present cross-sectional study aimed to investigate the mental health status and related factors of families of HCWs in Designated Hospitals in Ningbo, China.

**Method:** Family members of HCWs working in five designated hospitals in Ningbo, China were recruited between February 10th and 20th, 2020. Information on demographic variables, the COVID-19-related events in the lives, knowledge of COVID-19 and the working status of family members (that is, HCWs) was collected using online self-administered questionnaires. The mental health status were assessed using the Chinese version of Patient Health Questionnare-9 (PHQ-9) and Chinese version of Generalized Anxiety Disorder-7 (GAD-7). Multivariable logistic regression analyses were conducted to identify the main factors associated with the mental health conditions.

**Results:** A total of 822 participants completed questionnaires correctly. (response rate of 95.80%). The overall prevalence of GAD and depressive symptoms were 33.73%, and 29.35%, respectively. More times (hours) to focus on the COVID-19 (Odd ratio (OR)=1.215, 95% confidence interval (CI):1.061-1.391), family members (that is, HCWs) directly contact with confirmed or suspected COVID-19 patients (OR=1.477, 95%CI:1.069-2.040) were risk factors for GAD, while higher participants self-reported safety score for protective equipment of HCWs (OR=0.807, 95%CI:0.700-0.930) was a protective factor. More times (hours) to focus on the COVID-19 (OR=1.215, 95%CI:1.061-1.391), longer average working times per week for family members (that is, HCWs) (OR=1.017, 95%CI:1.005-1.029), being parents and other next of kin of HCWs were risk factors for depressive symptoms (OR=3.526, 95%CI:1.609-7.728 and OR=1.639, 95%CI:1.096-2.451, respectively). In addition, compared with participants who were HCWs, participants who were enterprise workers and were more likely to develop depressive symptoms(OR=1.750, 95%CI:1.104-2.776), while who were government employees or institutions employees were less likely to suffer depressive symptoms (OR=0.529, 95%CI:0.286-0.977).

**Conclusions:** Psychological responses to COVID-19 have been dramatic among family members of HCWs during the rising phase of the outbreak. Our findings provide strong evidence to pay more attention on the mental health status of this vulnerable but often unseen populations during COVID-19 epidemic.

## Introduction

Since December 2019, coronavirus disease 2019 (COVID-19) outbreak has occurred in Wuhan, Hubei Province, China, which has spread rapidly throughout China and even the world^1^. As of March 8, 2020, a total of 80,695 COVID-19 confirmed cases with 3,097 deaths had been reported in mainland China and the 21,110 confirmed cases with 413 deaths had been reported outside of China. In total, the global number of confirmed cases of COVID-19 has surpassed 100,000 in 93 countries/territories/areas^2,3^.

The first confirmed case of COVID-19 in Ningbo was reported on January 21, 2020^4^. As of March 8, 2020, a total of 157 COVID-19 cases have been confirmed in Ningbo^5^. In order to prevent and control the epidemic of COVID-19, the local government has set up fifteen designated-hospitals, of which five have treated for confirmed or suspected COVID-19 patients as of February 28, 2020^6^.

Compelling evidence have suggested that infectious disease pandemics, including severe acute respiratory syndrome (SARS), middle east respiratory syndrome (MERS) and 2009 novel influenza A (H1N1), were associated with mental health problems among the general population^7-9^, HCWs^10-14^, patients^15-17^and family members of patients^18^.

The severe situation of COVID-19 is also causing HCWs’ mental health problems such as stress, anxiety and depressive symptoms in China^19^. One of the most important reasons is that many HCWs lacked contacting with families^19^. Meanwhile, they may also experience fear of contagion and spreading the virus to their families^20^. Based on the above research evidence, we have reason to speculate that families of HCWs may also suffer from similar psychological problems to HCWs during COVID-19 epidemic^21-23^. As a result, in the National Health Commission guidelines for emergency psychological crisis intervention for people affected by COVID-19, families of HCWs were ranked as the third priority group^24^.

To date, series of studies have explored the psychological impact of COVID-19 outbreak on HCWs^19,20,26,27^. However, epidemiological data on the psychological impact among families of HCWs for COVID-19 epidemic have been limited.

Therefore, we performed a online-based cross-sectional study to investigate the mental health status and its influencing factors among families of HCWs who directly or indirectly contacted with confirmed or suspected COVID-19 patients in Ningbo, China. We hope our study findings will provide data support for the targeted interventions to this often unseen populations to cope with psychological problems during COVID-19 epidemic^28^.

## Methods

### Study design and participants

A cross-sectional study was conducted in Ningbo in February 2020. To prevent the spread of COVID-19 through droplets or contact, we used a online-based survey via the WeChat-based survey programme Questionnaire Star to collected data. With the help of five designated hospitals that have treated for confirmed or suspected COVID-19 patients in Ningbo, a total of 882 family members of HCWs from these five designated hospitals were invited to participate in this study. All participants were informed of study procedure upon their recruitment.

The inclusion criteria of this study were (1) being the next of kin of HCWs from designated-hospitals for medical treatment of COVID-19 in Ningbo; (2) having access to the internet; Exclusion criteria were as follows: (1) self-reported history of neurological disorders, mental illness and other serious systemic disorders; (2) self-reported substance abuse; Participants who passed the initial self-screening phase were asked to complete the self-administrated questionnaires^25^.This study was approved by the Ethical Committee of Ningbo First Hospital (approval number: 2020-R042) and registered with the registry website *http://www.chictr.org* (registration number: ChiCTR2000030697). Signed informed consent was obtained online from all participants.

### Measures

The questionnaires consisted of demographics, the Chinese version of Patient Health Questionnare-9 (PHQ-9), the Chinese version of Generalized Anxiety Disorder-7 (GAD-7) and questionnaires that were developed specifically for this study, as there were no suitable scales available for measuring the factors related to families of HCWs during COVID-19 outbreak. The self-developed questionnaires included the working status of family members (that is, HCWs), the participants’ knowledge of COVID-19, and the COVID-19-related events in the lives of participants.

### Demographics

The demographic characteristics included age, gender, educational level, occupation and the kinship with HCWs. Occupation included the following four types: (1)HCWs; (2) enterprise workers; (3) government employees or institutions employees; (4) students; (5) others, which consisted of freelancers, retirees, social workers, and other relevant staffs. The kinship of HCWs included the following four types:(1)spouses; (2)children; (3)parents; (4) other next of kin

### Self-developed questionnaires

The working status of family members (that is, HCWs) included: (1)whether HCWs directly contact with confirmed or suspected COVID-19 patients; (2) the average working time (hours) per week for HCWs; (3) participants’ self-reported safety score for protective equipment of HCWs (The safety scores ranging from 1 to 5, with the higher score indicating a better protective effect); (4) the department of HCWs, which was categorized into five items: a. front-line departments included respiratory department, infection department, ICU, fever clinic and isolation ward; b. medical technology department (imaging department and laboratory department, etc.);c. nursing department; d. logistics department; e. other departments.

The participants’ knowledge of COVID-19 was assessed by answering five single topic selection related to COVID-19: (1)which of the following symptoms is not a common symptom of COVID-19, with possible response options being fever, stuffy nose and runny nose, fatigue, and dry cough; (2) how many days do people returning from Hubei province need to be quarantined and observed, with possible response options being ten days, twelve days, fourteen days and fifteen days; (3) which of the following masks can prevent COVID-19, with possible response options being activated carbon mask, cotton mask, sponge mask and medical surgical mask; (4) the known transmission routes of COVID-19 do not include which of the following, with possible response options being contact transmission, droplet transmission, soil transmission and aerosol transmission; (5)with regard to the disposal of discarded masks, what is incorrect about the following statement, with possible response options being throw it away at any time when you run out of it, masks worn by people with fever need to be disinfected, sealed and discarded, wash your hands immediately after handling the mask and discarded masks should be discarded into hazardous trash cans. Of the above five questions, one point for each correct answer and no points for incorrect answers. A total score was calculated by summing points for each of the five questions, ranging from 0 to 5, with the higher score indicating a better knowledge of COVID-19.

The COVID-19-related events in the lives of participants included: (1) whether there had been confirmed COVID-19 cases in families or friends; (2) whether there had been suspected COVID-19 cases in families or friends; (3) times to focus on COVID-19, which was measured by the average hour spent focusing on the COVID-19 information every day.

### Patient Health Questionnare-9

We employed the Chinese version of PHQ-9 to assess the depressive symptoms of families of HCWs. PHQ-9 is a self-report, 9-item measure to assess depression severity. Participants rated each item in accordance with the frequency of symptoms over the past two weeks, on a 4-point scale, from 0 (not at all) to 3 (nearly every day). Total scores range from 0 to 27, with higher scores indicting greater severity of depressive symptoms^29^. The PHQ-9 has been widely used in China^30,31^.The good internal consistency reliability (Cronbach’s alpha coefficients ranged from 0.84 to 0.86) of the Chinese version of PHQ-9 has been demonstrated^32-34^. In the present study, Cronbach’s _α_ = 0.81. The depressive symptom was defined as a total score of ≥ 5 points in the PHQ-9 according to the previous study during COVID-19 epidemic^25^.

### Generalized Anxiety Disorder-7

We employed the Chinese version of GAD-7 to assess the GAD of families of HCWs. GAD-7 is a self-report questionnaire that screening and measure severity of GAD. Participants rated seven items according to the frequency of symptoms in the past two weeks, on a 4-point scale, from 0 (not at all) to 3 (nearly every day). Total scores range from 0 to 21, with higher scores indicting greater severity of GAD^35^. The GAD-7 has also been widely used in China and the good internal consistency reliability (Cronbach’s alpha coefficients ranged from 0.84 to 0.88) of the Chinese version of GAD-7 has been confirmed^36-40^. In the present study, Cronbach’s _α_ = 0.84.The presence of anxiety was defined as a total score of ≥5 points in the GAD-7 according to the previous study during COVID-19 epidemic^25^.

### Statistical analysis

Categorical variables were described as percentages and were compared using chi-square tests. Continuous variables were described as mean±SD and were compared using ANOVA analyse. Variables with skewed distributions were log-transformed to normal distribution. Multivariable logistic regressions were used to assess the associations between the PHQ-9 and GAD-7 score and potential related factors. Statistical analysis were performed using SPSS 22.0 (SPSS Inc., Chicago, IL,USA). Two-sided p-value <0.05 was considered statistically significant.

## Results

### Demographic characteristics

Of the 882 participants who were recruited in this study, 845 completed the questionnaires correctly (response rate of 95.80%). Table 1 presents characteristics of participants.

**Table 1.** Sample characteristics and associations with generalized anxiety disorder and depressive symptoms

| Variables | n (%)/$\bar{x} \pm \sigma$ | GAD | | *p* | Depressive symptom | | *p* |
| --- | --- | --- | --- | --- | --- | --- | --- |
|  |  | (GAD-7 score) |  |  | (PHQ-9 score) |  |  |
|  |  | <5(n=560) | ≥5(n=285) |  | <5(n=597) | ≥5(n=248) |  |
| Demographics |  |  |  |  |  |  |  |
| Gender |  |  |  | 0.042 |  |  | 0.012 |
| Male | 445(52.66) | 309(55.18) | 136(47.72) |  | 331(55.44) | 114(45.97) |  |
| Female | 400(47.34) | 251(44.82) | 149(52.28) |  | 266(44.56) | 134(54.03) |  |
| Age(years) | 37.98±9.39 | 37.43±9.23 | 39.07±9.62 | 0.016 | 37.62±9.29 | 38.83±9.59 | 0.088 |
| Educational level |  |  |  | 0.877 |  |  | 0.785 |
| Secondary school or below | 40(4.73) | 26(4.64) | 14(4.91) |  | 29(4.86) | 11(4.44) |  |
| High school | 63(7.46) | 43(7.68) | 20(7.02) |  | 41(6.87) | 22(8.87) |  |
| Junior college or bachelor | 650(76.92) | 427(76.25) | 223(78.25) |  | 462(77.39) | 188(75.81) |  |
| Master or above | 92(10.89) | 64(11.43) | 28(9.82) |  | 65(10.89) | 27(10.89) |  |
| Occupation |  |  |  | 0.340 |  |  | 0.007 |
| HCWs | 397(46.98) | 259(46.25) | 138(48.42) |  | 279(46.73) | 118(47.58) |  |
| Enterprise workers | 191(22.60) | 122(21.79) | 69(24.21) |  | 128(21.44) | 63(25.40) |  |
| Government employees or<br>institutions employees | 102(12.07) | 74(13.21) | 28(9.82) |  | 84(14.07) | 18(7.26) |  |
| Students | 21(2.49) | 17(3.04) | 4(1.40) |  | 19(3.18) | 2(0.81) |  |
| Others | 134(15.86) | 88(15.71) | 46(16.14) |  | 87(14.57) | 47(18.95) |  |
| Kinship of HCWs |  |  |  | 0.158 |  |  | 0.001 |
| Spouses | 553(65.44) | 365(65.18) | 188(65.96) |  | 405(67.84) | 148(59.68) |  |
| Children | 40(4.73) | 29(5.18) | 11(3.86) |  | 34(5.70) | 6(2.42) |  |
| Parents | 49(5.80) | 26(4.64) | 23(8.07) |  | 26(4.36) | 23(9.27) |  |
| Other next of kin | 203(24.02) | 140(25.00) | 63(22.11) |  | 132(22.11) | 71(28.63) |  |
| **The COVID-19-related events in the lives of participants** |  |  |  |  |  |  |  |
| **Whether there had been confirmed COVID-19 cases in families or friends** |  |  |  | 0.264 |  |  | 0.879 |
| Yes | 3(0.36) | 1(0.18) | 2(0.70) |  | 2(0.34) | 1(0.40) |  |
| No | 842(99.64) | 559(99.82) | 283(99.30) |  | 595(99.66) | 247(99.60) |  |
| **Whether there had been suspected COVID-19 cases in families or friends** |  |  |  | 0.186 |  |  | 0.501 |
| Yes | 70(8.28) | 41(7.32) | 29(10.18) |  | 47(7.87) | 23(9.27) |  |
| No | 775(91.72) | 519(92.68) | 256(89.82) |  | 550(92.13) | 225(90.73) |  |
| **Times to focus on COVID-19 per day (hours)** |  |  |  | **0.006** |  |  | **0.018** |
| <1 | 223(26.39) | 160(28.57) | 63(22.11) |  | 160(26.80) | 63(25.40) |  |
| 1-2 | 298(35.27) | 203(36.25) | 95(33.33) |  | 222(37.19) | 76(30.65) |  |
| 2-3 | 112(13.25) | 77(13.75) | 35(12.28) |  | 83(13.99) | 29(11.69) |  |
| >3 | 212(25.09) | 120(21.43) | 92(32.28) |  | 132(22.11) | 80(32.26) |  |
| **Knowledge of COVID-19** | 4.45±0.68 | 4.46±0.66 | 4.44±0.70 | 0.653 | 4.47±0.66 | 4.42±0.72 | 0.348 |
| **The working status of family members (that is, HCWs)** |  |  |  |  |  |  |  |
| **The department of HCWs** |  |  |  | 0.981 |  |  | 0.458 |
| Front-line departments | 131(15.5) | 86(15.36) | 45(15.79) |  | 92(15.41) | 39(15.73) |  |
| Medical technology department | 69(8.17) | 46(8.21) | 23(8.07) |  | 50(8.38) | 19(7.66) |  |
| Nursing department | 287(33.96) | 192(34.29) | 95(33.33) |  | 213(35.68) | 74(29.84) |  |
| Logistics department | 43(5.09) | 30(5.36) | 13(4.56) |  | 28(4.69) | 15(6.05) |  |
| Other departments | 315(37.28) | 206(36.79) | 109(38.25) |  | 214(35.85) | 101(40.73) |  |
| **Whether HCWs directly contact with confirmed or suspected VOVID-19 patients** |  |  |  | **0.019** |  |  | 0.781 |
| Yes | 406(48.05) | 253(45.18) | 153(53.68) |  | 285(47.74) | 121(48.79) |  |
| No | 439(51.95) | 307(54.82) | 132(46.32) |  | 312(52.26) | 127(51.21) |  |
| **The average working time per week for HCWs (hours)** | 41.44±13.07 | 40.73±12.39 | 42.83±14.23 | **0.034** | 40.70±12.16 | 43.21±12.93 | **0.020** |
| **Participants' self-reported safety score for protective equipment of HCWs** | 3.78±1.08 | 3.85±1.07 | 3.62±1.09 | **0.004** | 3.81±1.10 | 3.70±1.03 | 0.178 |

Abbreviations: n=number; GAD=Generalized Anxiety Disorder; PHQ= Patient Health Questionnaire; HCWs= health care workers; COVID-19=2019 Corona Virus Disease.
Notes: Numbers in bold indicate statistical significance at the 5% level.

The mean age of participants was 37.98±9.39 years. Among these samples, 52.66% were male, nearly a half (46.98%) were also HCWs, 65.44% were in a spousal relationship with the HCWs, and most of participants (approximately 87.81%) had a Junior college or bachelor or above education.

Only 0.36% and 8.28% of participants had confirmed and suspected COVID-19 cases in families or friends, respectively. 25.09% of participants focused on the COVID-19 for 3 hours or more every day. The mean score of participants’ knowledge of COVID-19 was 4.45±0.68. Almost one-sixth (15.5%) of participants’ family members (that is, HCWs) worked in the Front-line departments, nearly a half (48.05%) have to directly contact with confirmed or suspected VOVID-19 patients, the mean working time per week for HCWs was 41.44±13.07 hours, and the mean points of participants’ self-reported safety score for protective equipment of HCWs was 3.78±1.08.

### Prevalence of GAD and depressive symptoms among families of HCWs during COVID-19 epidemic stratified by variables

The prevalence of GAD and depressive symptoms among families of HCWs stratified by all the variables were also shown in Table 1. The overall prevalence of GAD and depressive symptoms were 33.73%, and 29.35%, respectively. There was statistically significant difference in the prevalence of GAD by gender, age, times to focus on COVID-19 per day (hours), whether HCWs directly contact with confirmed or suspected VOVID-19 patients, the average working time per week for HCWs (hours), and self-reported safety score for protective equipment of HCWs. There was statistically significant difference in the prevalence of depressive symptoms by gender, age, occupation, kinship of HCWs, times to focus on COVID-19 per day (hours), and the average working time per week for HCWs (hours).

### Variables associated with GAD and depressive symptoms among families of HCWs during COVID-19 epidemic

The associations of potential influence factors with GAD and depressive symptoms among families of HCWs during COVID-19 epidemic were presented in Table 2.

**Table 2.**
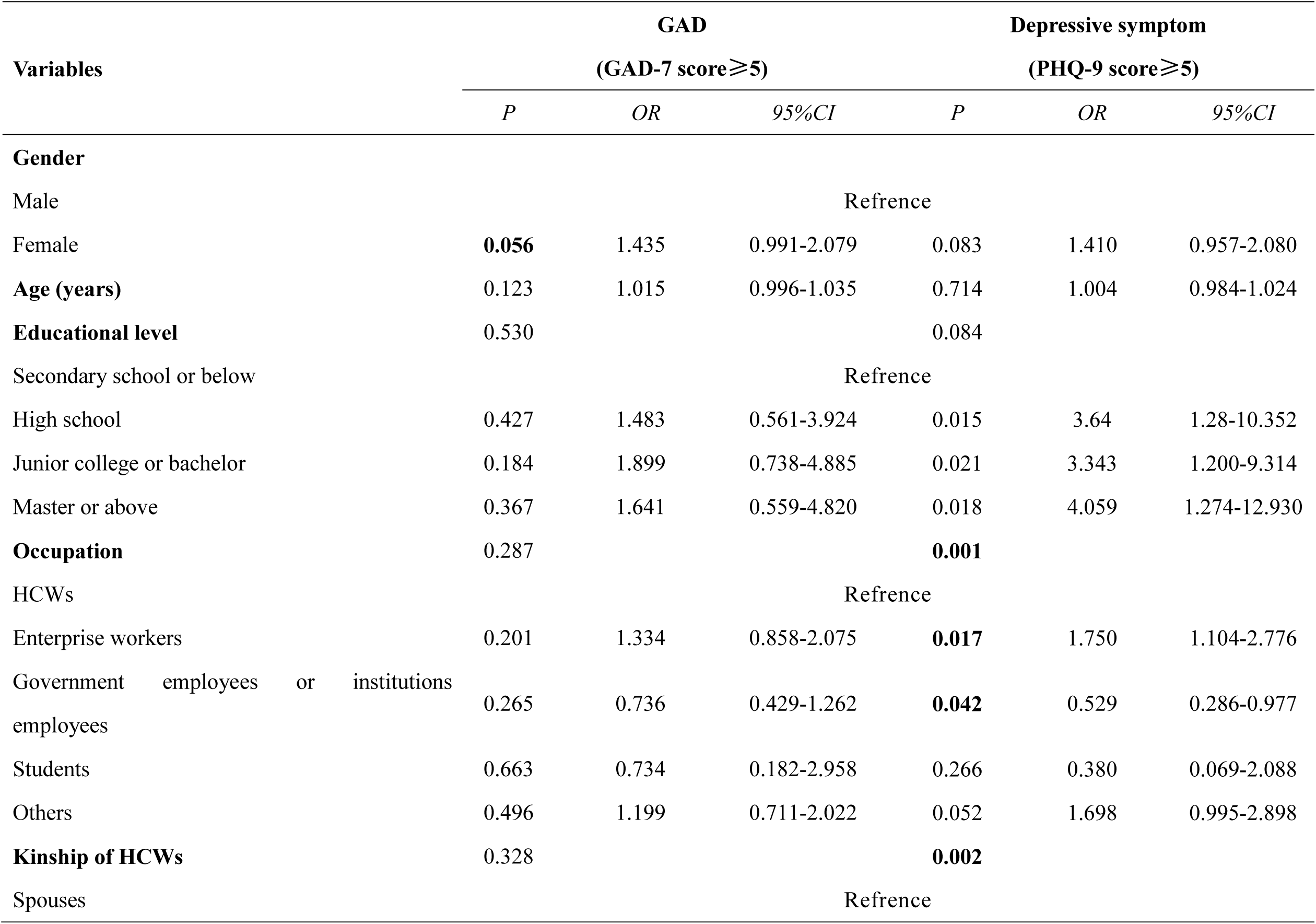
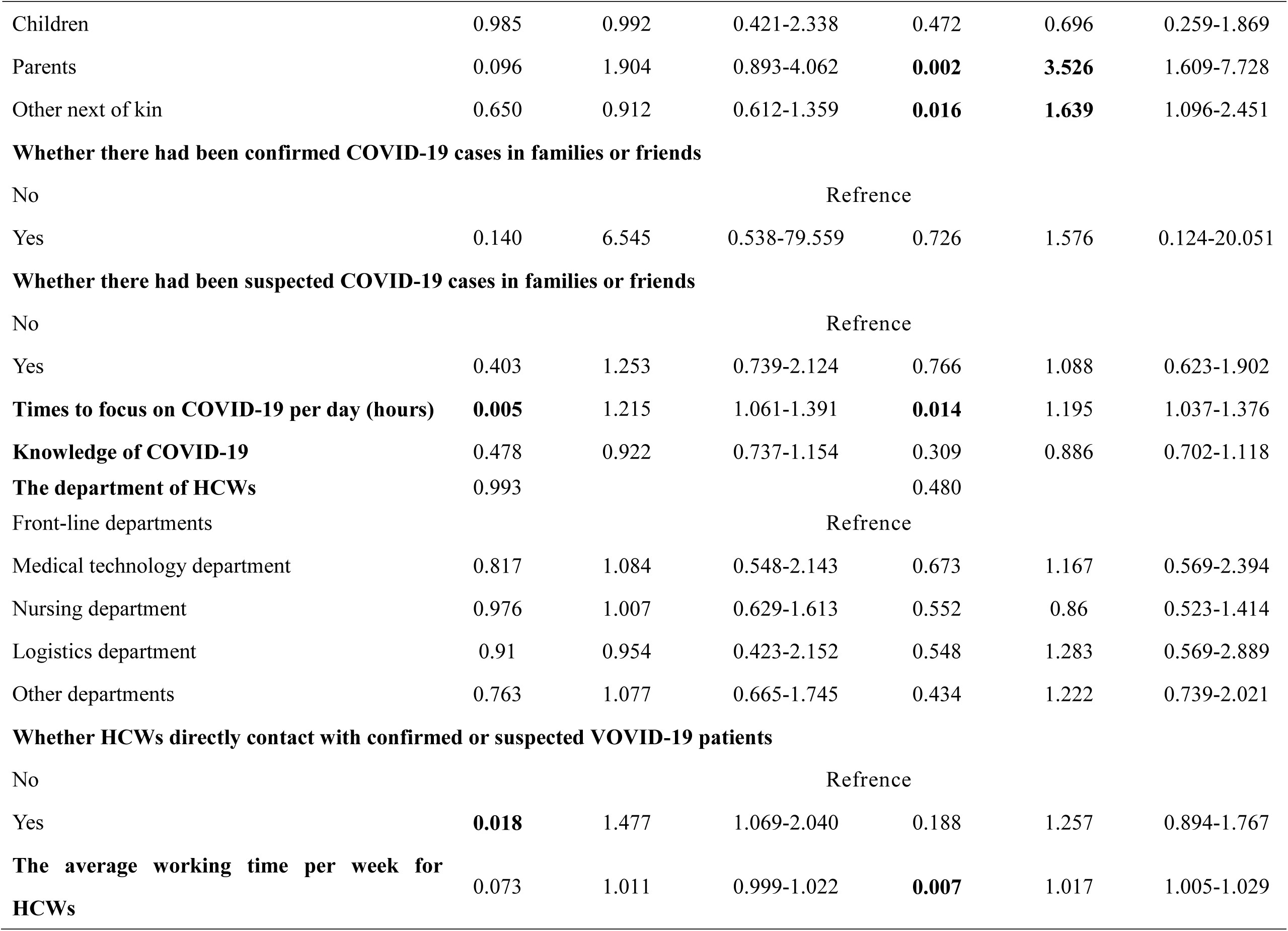
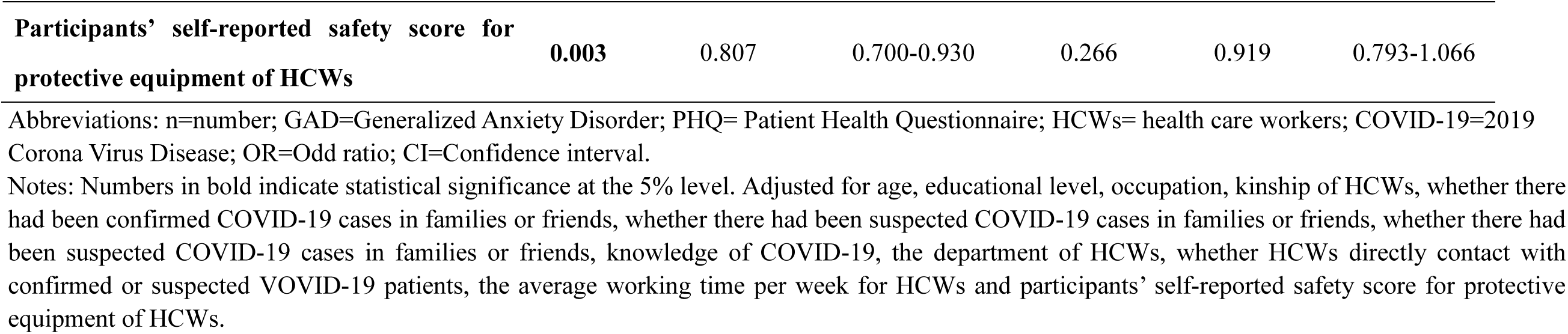
Factors associated with generalized anxiety disorder and depressive symptoms

| Variables | GAD |  |  | Depressive symptom |  |  |
| --- | --- | --- | --- | --- | --- | --- |
| | (GAD-7 score $\geq 5$) | | | (PHQ-9 score $\geq 5$) | | |
|  | *P* | *OR* | *95%CI* | *P* | *OR* | *95%CI* |
| **Gender** |  |  |  |  |  |  |
| Male |  |  | Reference |  |  |  |
| Female | **0.056** | 1.435 | 0.991-2.079 | 0.083 | 1.410 | 0.957-2.080 |
| **Age (years)** | 0.123 | 1.015 | 0.996-1.035 | 0.714 | 1.004 | 0.984-1.024 |
| **Educational level** | 0.530 |  |  | 0.084 |  |  |
| Secondary school or below |  |  | Reference |  |  |  |
| High school | 0.427 | 1.483 | 0.561-3.924 | 0.015 | 3.64 | 1.28-10.352 |
| Junior college or bachelor | 0.184 | 1.899 | 0.738-4.885 | 0.021 | 3.343 | 1.200-9.314 |
| Master or above | 0.367 | 1.641 | 0.559-4.820 | 0.018 | 4.059 | 1.274-12.930 |
| **Occupation** | 0.287 |  |  | **0.001** |  |  |
| HCWs |  |  | Reference |  |  |  |
| Enterprise workers | 0.201 | 1.334 | 0.858-2.075 | **0.017** | 1.750 | 1.104-2.776 |
| Government employees or institutions employees | 0.265 | 0.736 | 0.429-1.262 | **0.042** | 0.529 | 0.286-0.977 |
| Students | 0.663 | 0.734 | 0.182-2.958 | 0.266 | 0.380 | 0.069-2.088 |
| Others | 0.496 | 1.199 | 0.711-2.022 | 0.052 | 1.698 | 0.995-2.898 |
| **Kinship of HCWs** | 0.328 |  |  | **0.002** |  |  |
| Spouses |  |  | Reference |  |  |  |
| Children | 0.985 | 0.992 | 0.421-2.338 | 0.472 | 0.696 | 0.259-1.869 |
| Parents | 0.096 | 1.904 | 0.893-4.062 | **0.002** | **3.526** | 1.609-7.728 |
| Other next of kin | 0.650 | 0.912 | 0.612-1.359 | **0.016** | **1.639** | 1.096-2.451 |
| **Whether there had been confirmed COVID-19 cases in families or friends** |  |  |  |  |  |  |
| No |  |  |  | Refrence |  |  |
| Yes | 0.140 | 6.545 | 0.538-79.559 | 0.726 | 1.576 | 0.124-20.051 |
| **Whether there had been suspected COVID-19 cases in families or friends** |  |  |  |  |  |  |
| No |  |  |  | Refrence |  |  |
| Yes | 0.403 | 1.253 | 0.739-2.124 | 0.766 | 1.088 | 0.623-1.902 |
| **Times to focus on COVID-19 per day (hours)** | **0.005** | 1.215 | 1.061-1.391 | **0.014** | 1.195 | 1.037-1.376 |
| **Knowledge of COVID-19** | 0.478 | 0.922 | 0.737-1.154 | 0.309 | 0.886 | 0.702-1.118 |
| **The department of HCWs** | 0.993 |  |  | 0.480 |  |  |
| Front-line departments |  |  |  | Refrence |  |  |
| Medical technology department | 0.817 | 1.084 | 0.548-2.143 | 0.673 | 1.167 | 0.569-2.394 |
| Nursing department | 0.976 | 1.007 | 0.629-1.613 | 0.552 | 0.86 | 0.523-1.414 |
| Logistics department | 0.91 | 0.954 | 0.423-2.152 | 0.548 | 1.283 | 0.569-2.889 |
| Other departments | 0.763 | 1.077 | 0.665-1.745 | 0.434 | 1.222 | 0.739-2.021 |
| **Whether HCWs directly contact with confirmed or suspected COVID-19 patients** |  |  |  |  |  |  |
| No |  |  |  | Refrence |  |  |
| Yes | **0.018** | 1.477 | 1.069-2.040 | 0.188 | 1.257 | 0.894-1.767 |
| **The average working time per week for HCWs** | 0.073 | 1.011 | 0.999-1.022 | **0.007** | 1.017 | 1.005-1.029 |
| **Participants' self-reported safety score for protective equipment of HCWs** | **0.003** | 0.807 | 0.700-0.930 | 0.266 | 0.919 | 0.793-1.066 |

Abbreviations: n=number; GAD=Generalized Anxiety Disorder; PHQ= Patient Health Questionnaire; HCWs= health care workers; COVID-19=2019 Corona Virus Disease; OR=Odd ratio; CI=Confidence interval.
Notes: Numbers in bold indicate statistical significance at the 5% level. Adjusted for age, educational level, occupation, kinship of HCWs, whether there had been confirmed COVID-19 cases in families or friends, whether there had been suspected COVID-19 cases in families or friends, whether there had been suspected COVID-19 cases in families or friends, knowledge of COVID-19, the department of HCWs, whether HCWs directly contact with confirmed or suspected COVID-19 patients, the average working time per week for HCWs and participants' self-reported safety score for protective equipment of HCWs.

Multiple logistic regression analysis revealed that the participants who spent more times (hours) to focus on the COVID-19 (Odd ratio (OR)=1.215, 95% confidence interval (CI):1.061-1.391) and whose family members (that is, HCWs) directly contact with confirmed or suspected COVID-19 patients (OR=1.477, 95%CI:1.069-2.040) were significantly more likely to develop GAD, while higher participants’ self-reported safety score for protective equipment of HCWs (OR=0.807, 95%CI:0.700-0.930) was a significantly protective factor for participants to suffer GAD. In addition, female participants were marginally significantly more likely to have GAD (OR=1.435, 95%CI:0.991-2.079) than males.

Multiple logistic regression analysis also demonstrated that more times (hours) to focus on the COVID-19 (OR=1.215, 95%CI:1.061-1.391) and longer average working times per week for HCWs (OR=1.017, 95%CI:1.005-1.029) were significantly associated with higher risks of depressive symptoms among participants. Compared to participants who were HCWs, enterprise workers were significantly more likely to develop depressive symptoms (OR=1.750, 95%CI:1.104-2.776), while government employees or institutions employees were significantly less likely to have depressive symptoms (OR=0.529, 95%CI:0.286-0.977). Compared to participants who being a spouse of HCWs, parents and other next of kin were significantly much likely to develop depressive symptoms (OR=3.526, 95%CI:1.609-7.728 and OR=1.639, 95%CI:1.096-2.451, respectively)

## Discussion

This online-based cross-sectional study has provided evidence of a high prevalence of GAD and depressive symptoms among family members of HCWs in designated hospitals in Ningbo, China during COVID-19 epidemic. Therefore, as with HCWs, the mental health status of their families needs urgent attention. To the best of our knowledge, this is the first study to explore the mental health problems and its related factors among family members of HCWs during COVID-19 outbreak, and even the first study to investigate this issue during any infectious epidemic.

We note that 33.73% and 29.35% of family members of HCWs reported GAD and depressive symptoms, respectively. The prevalence of GAD and depressive symptoms were both much higher than the levels reported among general Chinese populations^41-44^. In addition, The prevalence of GAD and depressive symptoms were both lower than levels reported for a total of 1563 medical staff in a previous study using the same assessment instrument and cut-off score as this study (44.7% and 50.7%)^26^. It worth nothing that this study was conducted in the early phase of COVID-19 epidemic when most of Chinese HCWs had been facing the most severe situation, which is causing extreme psychological response^19^. Comparing our current study data with similar studies conducted at the same designed-hospitals it was interesting to note that, family members of HCWs were more likely to develop GAD and depressive symptoms than HCWs (28.8% and 23.9%, respectively). Although one needs to be cautious when comparing data from different studies using inconsistent time frames and medical conditions, this finding does demonstrate, to some extent, that an extreme psychological impact in association with COVID-19 epidemic, also in family members of HCWs.

Previous studies have demonstrated that hospital-related transmission was suspected to be the possible mechanism of infection for affected HCWs^45^. Thus, it is easy to understand that family members (that is, HCWs) directly contact with confirmed or suspected COVID-19 patients was associated with higher risk for GAD among participants, because they excessively concerned that their families might be infected or even die. This is consistent with the results of a previous study representing that family members of H1N1 patients suffer anxiety^18^.

Moreover, without proper personal protective equipment, COVID-19 may endanger HCWs^46^.As a result, it is possibly understandable that participants who report a higher safety score for protective equipment of HCWs would believe that their families were better protected, and therefore they were less likely to develop GAD.

Our findings demonstrated that longer average working times per week for HCWs was significantly associated with a higher risk of depressive symptoms among participants. Longer working times means that HCWs have to spent more time on contacting with confirmed or suspected COVID-19 patients, which may increase their chances of being infected with COVID-19^45^, and thus have a impact of depressive symptoms on their family members^18^. Longer working times would also occupy more family times of HCWs, and therefore may cause work-family conflict between HCWs and their family members, resulting in depression among both sides in the process of coping with this conflict^47^.

It is interesting to note that, compared to participants who were HCWs, enterprise workers were more likely to develop depressive symptoms (OR=1.750, 95%CI:1.104-2.776). Most enterprise workers have no medical background, and thus lack sufficient cognitive ability to raise awareness of COVID-19 outbreak. Their psychological endurance is lacking during a pandemic, and therefore may develop depressive symptom^48^. In addition, many enterprises were forced to shut down their work and production during the epidemic in China, resulting in the loss of income for their employees. However, income reduction was the predicting factor with the highest correlation of depression among enterprise workers, according to the results of a previous study during SARS in 2003^49^. Government employees or institutions employees were less likely to have depressive symptoms, which was less documented in the literature. It is likely that these populations were more likely to have access to a transparent announcement of epidemic information, so one possible explanation is that they do not have to suffer a feeling of uncertainty, which is a known risk factor for depression during a infectious epidemic^51^. Moreover, compared with HCWs who always suffer depression during the infectious epidemic, including COVID-19 outbreak^15,19,26,51^, most of government employees or institutions employees stayed away from confirmed or suspected COVID-19 patients, and therefore have lower risk of depressive symptoms than HCWs.

Meanwhile, similar to the results of a previous study during H1N1 in 2009, higher depression was noted for those in non-spousal relationships with HCWs, i.e. in our sample these were parents and other next of kin. Considering the majority of parents of HCWs were already elderly, one possible explanation is that rapid transmission of COVID-19 and high death rate have exacerbated the risk of mental health problems and worsen existing psychiatric symptoms among older adults^52^. We also speculate that most of parents and other next of kin of HCWs have no medical background, and therefore have a more extreme psychological response to COVID-19 epidemic, as we have discussed before^48^.

The only factor significantly associated with both GAD and depressive symptoms was times focus on COVID-19, which is consistent with findings in the general populations during COVID-19 outbreak^51^. Under this period of COVID-19 outbreak, most of the general populations, including family members of HCWs, stayed at home for isolation and faced with a great deal of information, as the government run national messaging campaigns that constantly emphasizing the dangers of COVID-19, especially on affected HCWs. Thus, family members of HCWs gained more time to gather information of HCWs who treated for COVID-19 patients, through the Internet and media^53^, for example on WeChat, which has spread the psychological influences, such as GAD and depressive symptoms, among the public more widely^20,54^. Moreover, the expression of this psychological reaction may be related to the normal protective response of the human body to the pressure of the epidemic, which also occurred in SARS outbreak in 2003^55^.

In reviewing the results of this study, there is evidence that insufficient and inadequate attention was paid to family members of HCWs during COVID-19 epidemic in China. Several appropriate interventions to alleviate GAD and depressive symptoms among this vulnerable population are recommended as follows: First, health policy makers and stakeholders should collaborate to provide high-quality, timely crisis psychological services to families of HCWs. Online psychological self-help intervention systems, including online cognitive behavioural therapy for depression and anxiety (eg, on WeChat), would be appropriate for families of HCWs^25,54^. Second, providing suitable protective equipment, work schedule, and accommodation to HCWs would benefit family members who were concerned about HCWs being infected. Third, according to the findings of previous studies^21,27^, social support appeared as a protective effect on mental health problems between HCWs and their families. Thus, we strongly recommend both sides to take the initiative to communicate with each other to show their support. Last but not least, government’s propaganda strategies should be well-organized and effective^56^. The provision of reliable and transparent epidemic information to family members of HCWs is essential to enhance their sense of control and self-efficacy, and therefore enable them to cope with the psychological impact of COVID-19 outbreaks^58^.

This study has several limitations. First, because we performed a cross-sectional study, our results do not show a causal relationship. Second, in order to prevent potential COVID-19 infection, a web-based survey was conducted, and therefore the sampling of our study was voluntary, resulting in possible selection bias. Third, our sample is not highly representative as the respondents were all from Ningbo city.

## Conclusion

The present study provided evidence of a major mental health burden of family members of HCWs worked in designated hospitals during COVID-19 epidemic in Ningbo, China, particularly in participants with more times to focus on COVID-19, family members (that is, HCWs) directly contact with confirmed or suspected COVID-19 patients, family members (that is, HCWs) have longer average working times per week, and who were in non-spousal relationships with HCWs. On the contrary, participants’ higher self-reported safety score for protective equipment of HCWs was a protective factor against psychological problems. Compared with participants who were HCWs, those who were enterprise workers were more likely to develop mental health problems, while who were government employees or institutions employees were less likely to report psychological response. In summary, we suggest that more attention should be paid on the mental health status of this vulnerable but often unseen populations in a infectious disease outbreak. In addition, our findings are important in enabling government to allocate health resources and offer appropriate treatments for family members of HCWs who suffer mental health problems during COVID-19 epidemic and any of the infectious disease outbreak in the future.

## Data Availability

The partial datasets generated or analysed during the current study are included in this preprint article and not publicly available due to have not formally published, but are available from the corresponding author on reasonable request.

## Acknowledgments

We thank all the family members of health care workers in our study. In addition, we express our heartfelt respect to all health care workers who are fighting against the COVID-19 epidemic.

## Authors’ contributions

YCY and ZZL drafted the manuscript, YCY, LMR and ZZL conceived and designed framework of this article, YCY, FQK, BBZ, YXJ and ZZL collected and analyzed the literature. All authors read and approved the final manuscript.

